# Confidence-based laboratory test reduction recommendation algorithm

**DOI:** 10.1101/2022.06.20.22276546

**Authors:** Tongtong Huang, Linda T. Li, Elmer V. Bernstam, Xiaoqian Jiang

**Affiliations:** School of Biomedical Informatics, UTHealth, Houston, TX, United States; Department of Pediatric Surgery, McGovern Medical School, UTHealth, Houston, TX, United States; Division of General Internal Medicine, Department of Internal Medicine, McGovern Medical School, UTHealth, Houston, TX, United States

## Abstract

Unnecessary laboratory tests present health risks and increase healthcare costs. We propose a new deep learning model to identify unnecessary hemoglobin (Hgb) tests for patients admitted to the hospital. Machine learning models might generate less reliable results due to noisy inputs containing low-quality information. We estimate prediction confidence to measure reliability of predicted results. Using a “select and predict” design philosophy, we aim to maximize prediction performance by selectively considering samples with high prediction confidence for recommendations. We use a conservative definition of unnecessary laboratory tests, which we define as stable and below the lower normal bound (LBNR). Our model accommodates irregularly sampled observational data to make full use of variable correlations (i.e., with other laboratory test values) and temporal dependencies (i.e., previous observations) in order to select candidates for training and prediction. Using data collected from a teaching hospital in Houston, our model achieves Hgb prediction performance with a normality AUC at 95.89% and a Hgb stability AUC at 95.94%, while recommending a reduction of 9.91% of Hgb tests that were deemed unnecessary.

## 1. Introduction

Laboratory over-utilization is common, especially in the United States[1,2]. Unnecessary blood draws waste resources and may harm patients by contributing to iatrogenic anemia[1]. Ideally, we want to minimize laboratory utilization while obtaining necessary information[1,3].

Recently, researchers have used machine learning to identify unnecessary laboratory tests. Most approaches leverage time series prediction methods to take advantage of previous patient information, such as autoregressive models[4], mixed-effect models[5], and traditional recurrent neural networks (RNN)[6,7]. Previous studies that identify unnecessary laboratory tests fall into two categories: *information gain*, and observability learning.

*Information gain* methods measure whether certain laboratory tests yield informative values. These approaches require an exact definition of unnecessary laboratory tests. Roy et al.[4] identify laboratory tests in the normal range as low yield laboratory tests, but they ignore events where the laboratory value changes from the normal to abnormal range; however, transitions such as these are likely to be clinically relevant events. Cismondi et al.[8] dichotomize laboratory tests into “information gain” or “no information gain” categories based on both normality and dropping levels. Aikens et al.[9] consider laboratory stability using both absolute value and standard deviation changes. Their models recommend eliminating laboratory tests that have little information gain. However, these algorithms lack confidence estimates to measure the reliability of predictions. Metrics that summarize the confidence of a prediction are important adjuncts for clinicians using the algorithm.

*Observability learning* approaches estimate the need for a lab to be checked in actual practice. A missing observation means that the laboratory test was not checked by the physician. Yu et al.[10,11] used a two-layer long-short term memory (LSTM) network with multiple fully-connected layers to estimate the observability of the next laboratory test. The min-max loss function increases prediction accuracy as the likelihood of observability (i.e., the necessity of conducting the next laboratory test) increases. Their first approach[10] has a narrow definition of the ground truth, which aims to predict the change rate for laboratory values. The second approach[11] extends the previous model by using multitask learning mechanisms to include predictions for abnormality (i.e., laboratory values beyond the normal range) and transition (i.e., laboratory values change from the normal to the abnormal range or vice versa). However, such recommendation algorithms are highly dependent on actual physician practices, which might be not optimal for laboratory test reduction algorithms. Relying on past practices might not be applicable to different or complicated clinical situations. Observability learning models, which work to predict the need for a test, are likely to recommend eliminating laboratory tests with a low prediction confidence. We believe that a better strategy may be to focus on eliminating laboratory tests that can be confidently predicted.

Our work is designed to reduce unnecessary hemoglobin (Hgb) tests based on the following assumptions. Sequential Hgb levels within the normal range imply that the patient is stable[12,13]. We further define that the Hgb test is “stable” when its immediate value does not change from normal to abnormal.

We explore approaches that enable our algorithm to make safe and confident predictions with features embedded in the algorithm. First, we introduce an *outcome-level* safety assurance, which uses a conservative (‘safe’) definition of unnecessary Hgb laboratory tests. We define ‘safe’ Hgb tests as ones that are predicted to be stable and remain normal. If the predicted normality is consistent with predicted stability, we will recommend eliminating the next Hgb test. Second, we will estimate the confidence of predicting Hgb test values as a *sample-level* safety assurance. We estimated the confidence for predicting Hgb at a specific time based on all available time-series data by introducing a selection predictor in the neural network architecture[14]. In training, this means the model will ignore some samples, whose inclusion would decrease the performance of the model. In testing, this means the selection predictor will not make recommendations if the selection confidence is below a certain threshold. Our main contribution is to integrate the confidence-based selection process for candidate samples into the training and testing phase, rather than estimating prediction confidence in a post-hoc manner.

## 2. Results

### 2.1 Dataset Description

Our data included 75,335 distinct inpatient encounters from a large urban hospital system in the southern United States. Because the model requires learning previous observations of laboratory data, we excluded 8,528 encounters with only one Hgb test. We also eliminated 4,328 encounters with systolic blood pressures less than 90 mmHg to focus on patients who were hemodynamically stable, leaving 62,479 unique encounters. This included 804 pediatric encounters (age < 18). The demographics are summarized in **S4 Table**. We divided the entire cohort into 80% training set and 20% test set. To estimate future Hgb values, we also included 11 other common laboratory tests:

- Electrolytes: Na (sodium), K (potassium), Cl (chloride), HCO3 (serum bicarbonate), Ca (total calcium), Mg (magnesium), PO4 (phosphate)
- Renal function: BUN (Blood Urea Nitrogen), Cr (creatinine)
- Complete Blood Count (CBC): Plt (platelet count), WBC (white blood count)

We included five relevant vital signs (i.e., peripheral pulse rate, diastolic blood pressure, systolic blood pressure, respiratory rate, and SpO2 percent), individual patient demographics (i.e., gender, race, and age), hours from the last observation, Hgb value changes, and missing value indicators.

### 2.2 Prediction Tasks

Considering patient data at {0,1,2,…, *t*}, where *t* denotes the timestep number, our confidence-based selection model has four predictive tasks: selection probability *p*_*t*_, Hgb normality *y*_*t*_, Hgb stability *z*_*t*_, and Hgb value *v*_*t*_. Specifically, predicting Hgb normality *y*_*t*_ and Hgb stability *z*_*t*_ are responsible for *outcome-level* safety assurance, and predicting selection probability *p*_*t*_ is responsible for *sample-level* safety assurance.

Selection probability *p*_*t*_ measures the confidence of predicting the next Hgb. Prediction confidence represents predictability, that is, reliability of Hgb predictions estimated by the model. Based on Hgb tests that are confidently predicted, our model can make more accurate predictions of normality and stability. The model selected Hgb candidates with high prediction confidences, whose *p*_*t*_ is greater than a selection threshold *τ*.

For the Hgb normality *y*_*t*_ task, if the predicted value was above the LBNR, the Hgb test was considered normal (i.e., *y*_*t*_ = 1). Otherwise, the Hgb test was cosnsidered abnormal (i.e., *y*_*t*_ = 0). We assume that Hgb results exceeding the upper bound of the normal range (UBNR) are relatively uncommon (e.g., polycythemia) or irrelevant (e.g., indicate dehydration or chronic hypoxia that are usually monitored using other modalities and were excluded from our analysis). Most clinical cases focus on dropping Hgb (e.g., bleeding), and only 0.37% of Hgb results in our samples were above the UBNR. In our population-driven model, instead of using a uniform LBNR over the entire population, we defined normal Hgb based on age and gender (see **Table 3**).

For the Hgb stability *z*_*t*_ task, a predicted value was considered stable (i.e., *z*_*t*_ = 1) if it does not change from normal to abnormal. For this task, we only considered a decreasing drop from normal to abnormal. Agreement between normality and stability predictions reinforces the confidence of the overall prediction.

The Hgb value *v*_*t*_ was an auxiliary task to improve primary prediction tasks – Hgb normality and Hgb stability. We expected that Hgb value predictions with confident normality and stability predictions would also closely approximate the actual values.

Given an encounter’s input features *X*, we defined that the ground truth of unnecessary Hgb tests satisfy the following condition {*X*| *y*_*t*_ = 1} ∩ {*X*| *p*_*t*_ = 1} ∩ {*X*|*p*_*t*_ > *τ*}. That is, among selected candidates, if the next Hgb value was predicted to be normal and stable, the model would recommend a ‘safe’ reduction.

### 2.3 Training and Evaluation Design

In the training stage, we conducted random mask corruption to transform some observations into zeros in order to simulate the impact of recommended lab test reduction (see **Methods**). In the test stage, we either converted omitted laboratory values to zeros (in reduction evaluation) or kept original laboratory values (in no-reduction evaluation).

- *Training protocol:* In the training stage, input features *X* were fed into the network model at a fixed length *T*. The prediction at every timestep *t* depends on the observations from all previous timesteps. We trained separate models at the target coverage rate *c* = {0.75,0.8,0.85,0.9, 0.95,1.0}, which reflects the expected proportion of laboratory candidates that are selected for possible reduction.
- *Reduction evaluation (practical setting):* In the test stage, we simulated dynamic laboratory reduction during the evaluation process. Starting at the initial timestep *t* = 0, the model was fed initial inputs *x*_*0*_. For the following timesteps *t* > 0, we conducted stepwise reduction estimation. If the model estimated the next Hgb to be normal and stable, which yielded a recommendation to omit that test, the next Hgb input would be set at a zero value. The reduction evaluation process iterated until the last timestep.
- *No-reduction evaluation (idealized setting):* Like the training stage, the second evaluation protocol performed a fixed evaluation process. At every timestep *t*, the model obtained all input features from the previous timesteps. In this process, we always used full observation to make future predictions, and no lab test was ever reduced in the prediction process. Model robustness can be estimated from the gap between *reduction* and *no-reduction* evaluations.

### 2.4 Label Prediction Performance

We report the results of several experiments that validate the effectiveness of our confidence-based model in predicting normality and stability labels. **Fig 1** represents the comparison results under reduction evaluation and no-reduction evaluation metrics. Our numerical results are represented in **Table 1** and **Table 2**.

**Fig 1.**
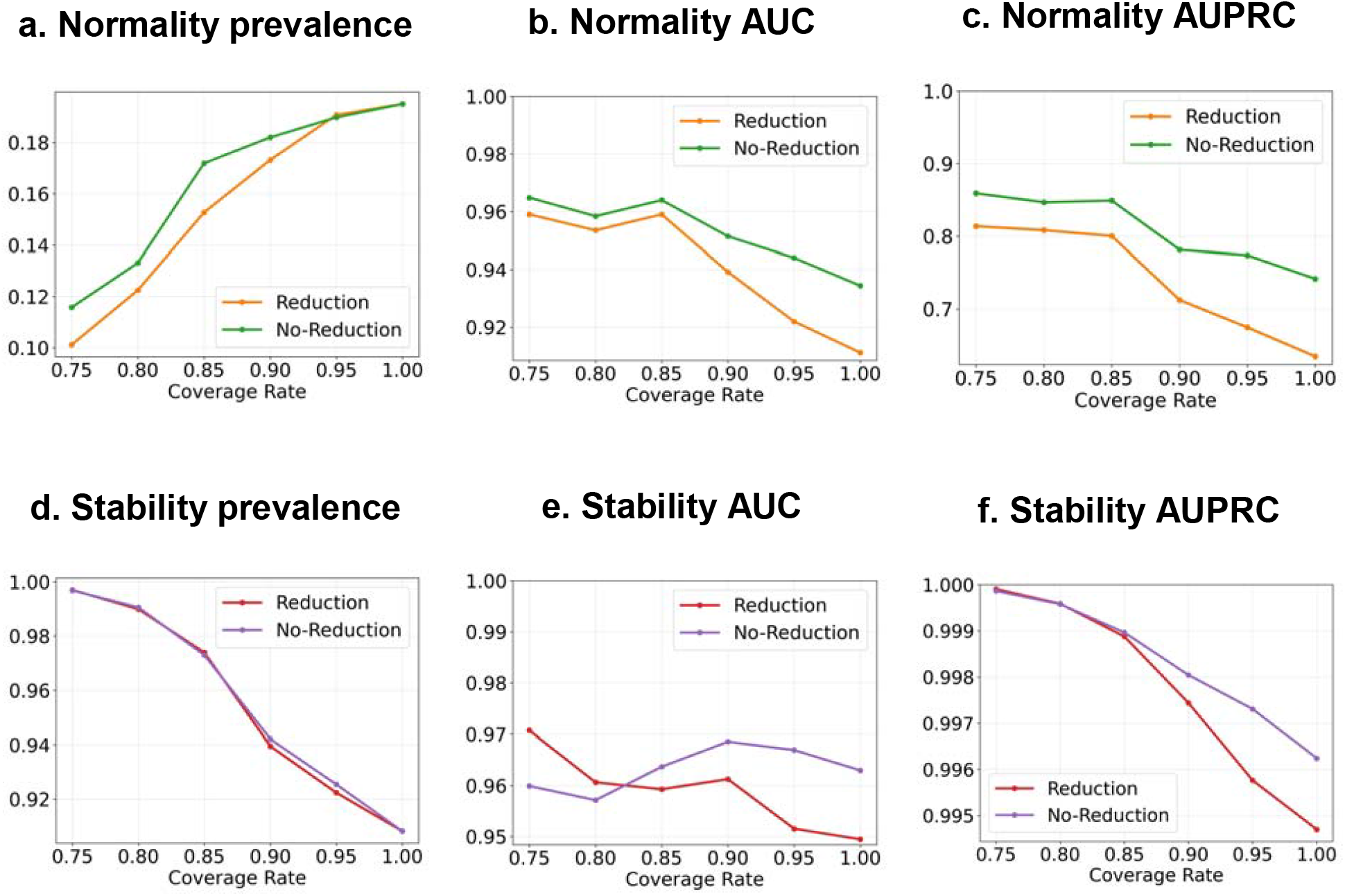
Model performance at multiple selection coverages under reduction and no-reduction evaluation. The selection threshold *τ* is 0.5. The coverage rate refers to the expected proportion of Hgb samples to constrict the model.

**Table 1.**
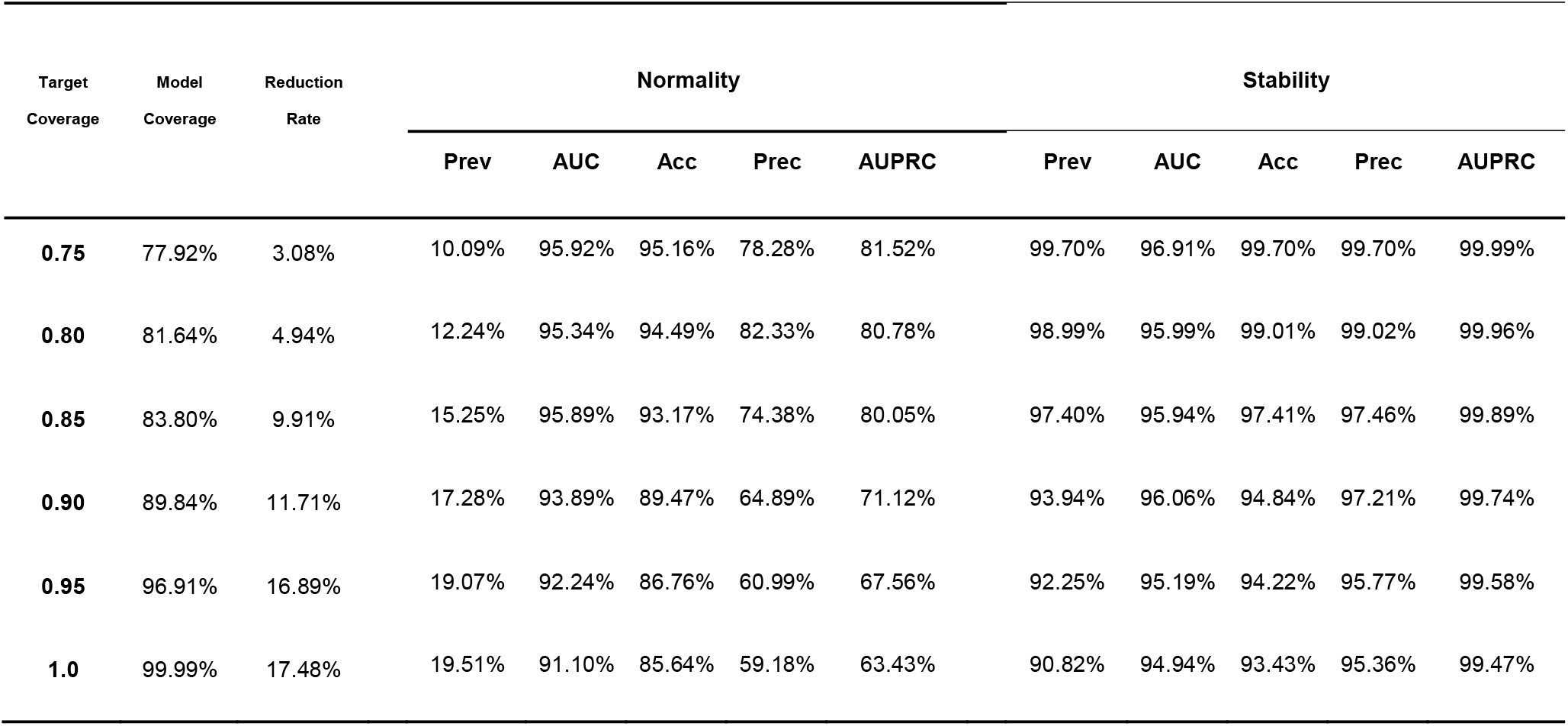
Model performance by selection coverages and reduction rate. The model coverage rate refers to the actual proportion of Hgb samples considered by the model; the reduction rate refers to the proportion of Hgb samples recommended for reduction among the entire Hgb test dataset; Prev indicates prevalence, which denotes the proportion of normal/stable Hgbs in selective candidates; AUC indicates area under the ROC curve; Acc indicates accuracy; Prec indicates precision; AUPRC indicates the precision-recall curve.

| Target Coverage | Model Coverage | Reduction Rate | Normality |  |  |  |  | Stability |  |  |  |  |
| --- | --- | --- | --- | --- | --- | --- | --- | --- | --- | --- | --- | --- |
|  |  |  | Prev | AUC | Acc | Prec | AUPRC | Prev | AUC | Acc | Prec | AUPRC |
| **0.75** | 77.92% | 3.08% | 10.09% | 95.92% | 95.16% | 78.28% | 81.52% | 99.70% | 96.91% | 99.70% | 99.70% | 99.99% |
| **0.80** | 81.64% | 4.94% | 12.24% | 95.34% | 94.49% | 82.33% | 80.78% | 98.99% | 95.99% | 99.01% | 99.02% | 99.96% |
| **0.85** | 83.80% | 9.91% | 15.25% | 95.89% | 93.17% | 74.38% | 80.05% | 97.40% | 95.94% | 97.41% | 97.46% | 99.89% |
| **0.90** | 89.84% | 11.71% | 17.28% | 93.89% | 89.47% | 64.89% | 71.12% | 93.94% | 96.06% | 94.84% | 97.21% | 99.74% |
| **0.95** | 96.91% | 16.89% | 19.07% | 92.24% | 86.76% | 60.99% | 67.56% | 92.25% | 95.19% | 94.22% | 95.77% | 99.58% |
| **1.0** | 99.99% | 17.48% | 19.51% | 91.10% | 85.64% | 59.18% | 63.43% | 90.82% | 94.94% | 93.43% | 95.36% | 99.47% |

**Table 2.** Model performance under selection coverages under no-reduction evaluation.

| Target Coverage | Model Coverage | Normality |  |  |  |  | Stability |  |  |  |  |
| --- | --- | --- | --- | --- | --- | --- | --- | --- | --- | --- | --- |
|  |  | Prev | AUC | Acc | Prec | AUPRC | Prev | AUC | Acc | Prec | AUPRC |
| **0.75** | 79.75% | 11.59% | 96.46% | 95.48% | 85.19% | 85.86% | 99.68% | 95.88% | 99.68% | 99.68% | 99.99% |
| **0.80** | 83.05% | 13.33% | 95.85% | 94.96% | 87.38% | 84.72% | 99.06% | 95.69% | 99.08% | 99.09% | 99.96% |
| **0.85** | 86.99% | 17.18% | 96.39% | 93.85% | 80.36% | 84.80% | 97.29% | 96.37% | 97.31% | 97.35% | 99.90% |
| **0.90** | 90.96% | 18.21% | 95.14% | 91.65% | 72.81% | 78.10% | 94.20% | 96.87% | 95.09% | 97.55% | 99.80% |
| **0.95** | 96.58% | 19.00% | 94.39% | 89.88% | 69.31% | 77.37% | 92.52% | 96.70% | 94.54% | 96.23% | 99.73% |
| **1.0** | 99.99% | 19.51% | 93.44% | 88.53% | 66.23% | 74.14% | 90.82% | 96.30% | 93.46% | 95.64% | 99.62% |

**Table 3.** Hgb normal range stratification table. LBNR means the lower bound of the normal range. UBNR means the upper bound of the normal range. Our experiments only considered Hgb LBNR to identify normality and stability.

| Age | Hgb LBNR (g/dL) | Hgb UBNR (g/dL) |
| --- | --- | --- |
| 6 months to 6 years | 10.5 | 14.5 |
| 7 - 12 years | 11.0 | 16.0 |
| Adult Women | 12.0 | 16.0 |
| Adult Men | 14.0 | 18.0 |

We set the default selection threshold *τ* = 0.5. The model selects a subset of Hgb samples with high prediction confidences and use *p*_*t*_ > 0.5 to make classifications, guided by the target coverage rate *c* (see **Method**). The selected samples are considered high-confidence candidates. A lower coverage rate means the model considers more high-confidence candidates for training and testing. But there is a tradeoff, as being too strict on prediction confidence would reduce selection size and hurt the model’s generalizability. “Model coverage rate” is the actual proportion of Hgb samples considered by the model. One can see that “model coverage rates” were close to “target coverage rates”, and they had less than 2% of differences in all settings, showing that the model is enforcing the target coverage rate.

Because the data distribution was skewed (i.e., approximately 19.5% of labs were normal and only 9.2% transitioned from normal to abnormal), we used AUC and AUPRC to model performance measures. **Fig 1** and **Table 1** show that all the selective models achieved normality AUCs at over 90%, and stability AUCs at over 94%, even in the extreme coverage case at *c* = 1. Note that even though we set *c* = 1, our model did not think all the Hgb predictions are necessarily good candidates for reduction (indeed, it considered 99% of Hgb samples were eligible for reduction recommendation).

As a good tradeoff between performance and reduction, we observed that the model reduced 9.91% Hgb tests at a coverage rate of 0.85. Most AUCs and AUPRCs decreased when the coverage rate increased. Note that stability AUCs did not drop at a steady rate because the proportion of stable samples was much larger than unstable ones, resulting in the mean probability bias favoring stable lab tests[15]. Overall, reduction evaluations achieved comparable performances over no-reduction evaluations. The results show that our model is robust in the practical setting when some input laboratory values are missing due to previous reductions.

We defined prevalence as the proportion of positive samples (i.e., normal and stable Hgbs) in the selected Hgb candidates. As the coverage rate increased, the normality prevalence increased linearly, and the stability prevalence decreased linearly, suggesting that the higher-confidence model tends to select more samples from the dominating class of labels (i.e., abnormal Hgbs and stable Hgbs) to obtain higher accuracy while recognizing minor labels (i.e., normal Hgbs and unstable Hgbs).

### 2.5 Value Prediction Performance

We evaluated the consistency between predicted values and predicted normality. The test was conducted under a reduction evaluation. Ideally, when the model estimates a Hgb sample to be normal, we expect its predicted value should also lie above the LBNR.

**Fig 2** (see numerical details in **S6 Table**) shows that the model predicted values that were consistent with normality predictions. Among Hgbs that were predicted to be normal, our model predicted >90% of corresponding values above the LBNR in a tolerable error of 3% at coverage rates <85%, and in a tolerable error of 5% at almost all the coverage rates. These results imply that our multitask learning framework is able to leverage the auxiliary task (value prediction) to support the primary prediction tasks (normality and stability prediction). Another observation is that the coverage rate has a large effect on recognizing normalities of predicted values. When the coverage rate is higher, the accuracy of normality prediction shows more variability at error rates 0-10%, suggesting that the low-confidence model has more value variability.

**Fig 2.**
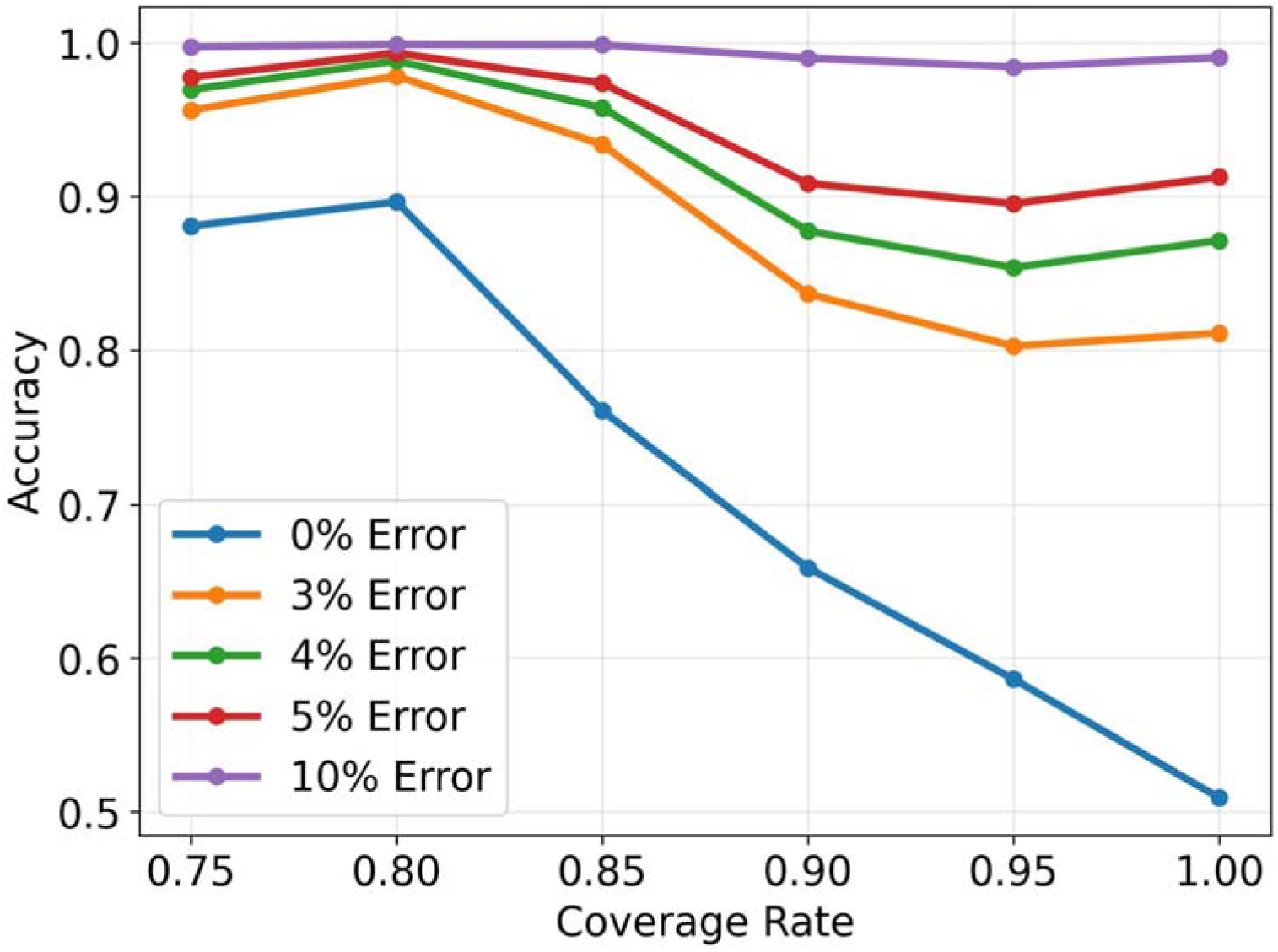
Consistency between predicted values and predicted normality. The objective was to measure the consistency between predicted values and predicted normalities. The “normality accuracy of predicted values” was defined as the percentage of predicted values with *f*_*V*_ (*x*_*t*_) > = *b*, where *b* is the value of the LBNR, on Hgb samples with *f*_*Y*_(*x*_*t*_) = 1. We considered a tolerable boundary to be m% lower than the LBNR for predicted normal Hgbs.

### 2.6 Selection Performance

The selection threshold *τ* was set at 0.5 during training. In the test phase, we evaluated the model using the default threshold *τ* = 0.5. In this section, we measured the influence of different selection thresholds *τ* ∈ [0.05,0.95] (with intervals at 0.05) on classification accuracy. The experiment was evaluated under laboratory reductions. We set the target coverage rate to 0.85. Our model accounted for 83.80% of Hgb candidates (**Fig 3**), using the default selection probability *τ* = 0.5.

**Fig 3.**
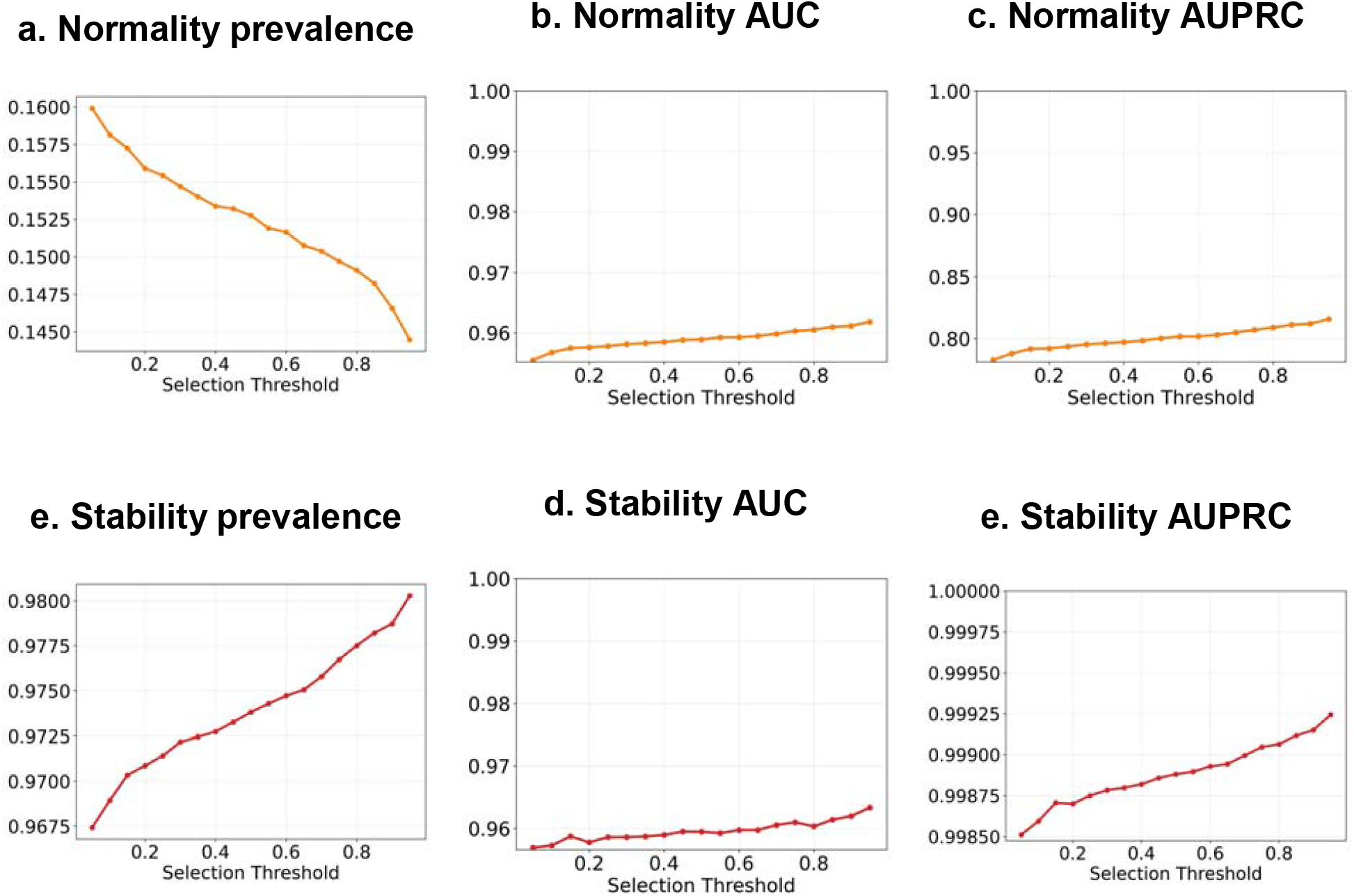
Model performances using different selection thresholds for reduction evaluation. The range of our selection threshold is *τ* ∈ [0.05,0.95] with intervals at 0.05.

As shown in **Fig 3**, although performance curves fluctuate at nearby selection thresholds, the trend remained increasing when the threshold value was increased. With a larger threshold, the model selected fewer normal Hgbs and more stable Hgbs, but obtained higher confidence in classifications. When the threshold *τ* is above 0.5, we achieved normality predictions over 95.8% AUC and 80.0% AUPRC, while stability predictions achieved more than 95.9% AUC and 99.8% AUPRC.

## 3. Discussion

We introduced a deep learning model with a selective framework to address the laboratory reduction problem. We conducted a case study on Hgb reduction. Based on the definition that unnecessary laboratory tests are the ones predicted to be normal and stable, we showed that our selective model achieved good predictive performance. Our major contribution is to offer safe recommendations for omitting unnecessary Hgb samples by jointly considering the confidence and prediction accuracy during training and testing. The idea was to select a proportion of Hgb candidates with high prediction confidence in estimating normality and stability. Our model automatically identified an appropriate balance between stability, normality, and prediction confidences, which achieved a performance of 95.89% normality AUC and 95.94% stability AUC with a potential to eliminate 9.91% of Hgb tests. In addition, when future Hgb tests were predicted as normal, our model predicted >90% of the corresponding predicted Hgb values within a tolerable error range of 3% at a 90% selection coverage, demonstrating its robustness. We also made some technical contributions in order to handle irregular time sequences in laboratory reduction by introducing a feed-forward attention function to capture the importance of every timestep. The ablation study results (see **S3 Table**) confirmed that both selective mechanism and attention-based LSTM layers contributed to the improvement of model performances.

Not all predictable laboratory tests are unnecessary in complicated clinical situations. To ensure usability, our model discovered unnecessary Hgb tests among predicted high-confidence candidates. The predicted low-confidence Hgb samples are not recommended for elimination, for which our model would acknowledge ‘I don’t know’ and let clinicians decide. Our model also offers a tunable parameter to control the confidence level for sample inclusion (i.e., expected selection coverage). Clinicians can choose a confidence level based on the context to receive recommendations for clinical decision making.

Nevertheless, our work still has limitations. First, the selection mechanism is computationally expensive because it needs to accommodate multiple objectives. Second, the optimal expected coverage rate that controls the model’s confidence might vary by different situations, and it is not directly interpretable as confidence intervals that are familiar to clinicians. Third, our study was internally validated for patients admitted to a single health institution. External validation studies are needed. Fourth, our model recommends reduction for an individual laboratory test. In clinical practice, Hgb tests are commonly ordered as a part of a complete blood count panel, which includes platelet count and white blood count. Our model currently does not account for bundled test reduction strategies because the laboratory panel information is missing in the dataset. Finally, the ground truth of unnecessary laboratory tests might not be perfect. Our model relies on standard of laboratory normal ranges, whereas some abnormal results may be predictable and stable (e.g., a clinically stable patient with stable anemia) and thus could be omitted. We will address these limitations in future research.

## 4. Methods

The workflow of the confidence-based candidate selection is demonstrated in **Fig 4**.

**Fig 4.**
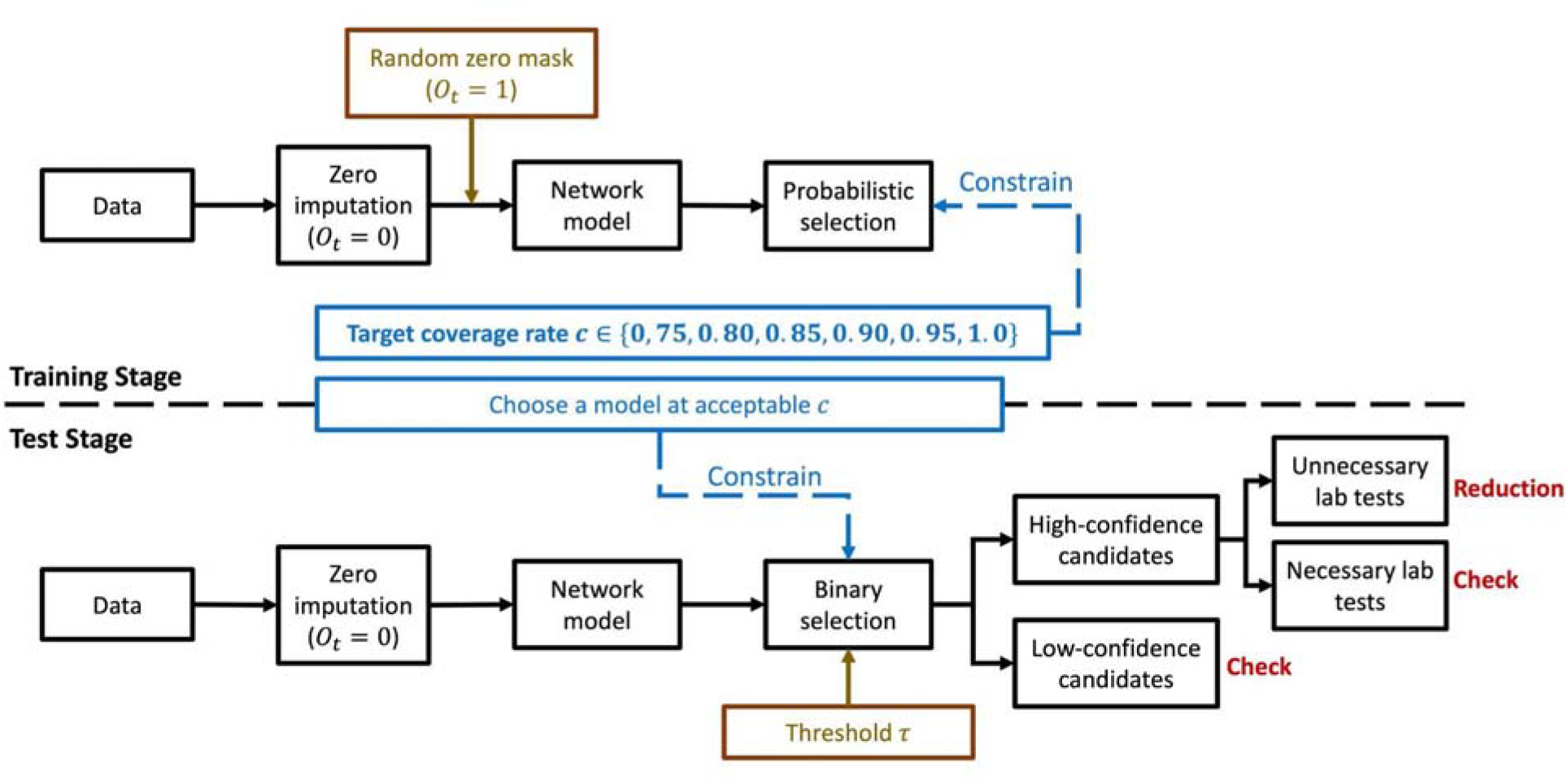
Block diagram of candidate selection. (1) Before processing the network model, we imputed zeros for missing features. (2) In the training stage, we inserted a random zero mask for existing laboratory values. The network model predicted selection probabilities for individual laboratory tests. Thus, the model ignored some samples, whose inclusions were considered to decrease performance. Each model was trained under one target coverage rate that constrained the actual proportion of selected laboratory tests. Intuitively, the lower coverage rate means that selections are more strict. (3) In the test stage, we chose a model at an acceptable coverage rate. A threshold *τ* was used to determine whether individual laboratory tests were selected.

The selected tests were considered high-confidence candidates. The model recommended canceling pending laboratory tests if predicted values satisfied two joint conditions: a. High-confidence; b. Unnecessary (i.e., predicted to be stable and remain normal).

### 4.1 Data Preprocessing

For each encounter, we organized laboratory test results into a consecutive sequence in temporal order. The laboratory tests conducted in the same hour were aggregated into a laboratory draw. If the same laboratory test appears more than one time at the encounter’s visit hour, we averaged the result values. All laboratory draws for each encounter start with the draw where the first Hgb result was recorded, and end with the draw where the last Hgb result was recorded. Finally, we capped the total length of laboratory draws to 30 timestamps (to make the parameter space tractable) based on the histogram of encounter visit times (i.e., the number of timestamped laboratory records for patients, see **S2 Fig**). To incorporate patient conditions, vital signs were also integrated with the laboratory draw by the event time. We calculated average values when more than one vital sign was reported during the same hour.

To better understand each patient, our model includes patient demographics, involving gender, race, and age information. The normal range varies depending on gender and age (**Table 3**). We encoded gender and race information as categorical variables. We also categorized patient ages into four groups based on their different normal ranges and genders.

### 4.2 Notation

**Table 4** explains the symbols used in our model. For each encounter, we had a multivariate time series data of length *T*. We gave symbols to represent important concepts: feature measurements *X*, Hgb normality labels *y*, Hgb stability labels *Z*, Hgb values *V*, and selection probability *P. X* represented a group of input features, including laboratory values, vital signs, Hgb value changes, demographics, time differences, and observation indicators. The observation indicator was denoted as *o*. It was used for handling missing values of the dataset. Hgb normality labels *Y*, Hgb stability labels *z*, and Hgb values *V* served as gold standards to measure our model’s prediction performance. To measure the confidence of predicting the next Hgb test, our model also predicted the selection probability *P*. As observations were not necessarily made at regular intervals, hence, the timestep *t* simply indexed the sequence of observations.

**Table 4.** Nomenclature table.

| Symbol | Explanation |
| --- | --- |
| $X$ | Input features (including laboratory values, vital signs, demographics, time differences in terms of hours, and observation indicators) |
| $O$ | Observation indicator. It is included in input features $X$. (If the laboratory sample is observed, $o_t = 1$; otherwise, $o_t = 0$ ) |
| $T$ | The total number of timesteps. In a time sequence, a record of a laboratory group occurs at one timestep. The maximum number of timesteps is 30 |
| $P$ | The selection probability (If the Hgb is selected, $p_t = 1$; otherwise, $p_t = 0$) |
| $Y$ | Normality label (If the Hgb value is above the LBNR, $y_t = 1$; otherwise, $y_t = 0$) |
| $Z$ | Stability label (If the Hgb value does not transit from normal to abnormal, $z_t = 1$; otherwise, $z_t = 0$) |
| $V$ | Hemoglobin value |
| $\tau$ | The selection threshold. Its default value is 0.5. (If predicted selection probability $p_t > \tau$, the corresponding Hgb sample is selected as a candidate for reduction; otherwise, the Hgb sample is not a candidate for reduction) |

When an encounter had observed data (i.e., a Hgb test drawn and resulted) at a timestep *t*, we denoted the observation indicator *o*_*t*_ = 1; Otherwise, we denoted *o*_*t*_ = 0. The normality *y*_*t*_, stability *z*_*t*_, values *v*_*t*_, and selection probabilities *p*_*t*_ represented the outcome realized at timestep *t*. We used a sigmoid function to output predictions for normality *Y*, stability *Z*, and selection probability *P* in a range of [0,1], and restricted predictions for *V* to be positive. To make a cutoff for classifications, we used a threshold for normality, stability, and selection probability. The default thresholds were 0.5. In the test stage, the selection threshold is adjustable, and we set values in the range of *τ* ∈ [0.05.0.95].

### 4.3 Feature Processing

This section introduces our methods to handle missing data and random mask corruption. We incorporated relational positional time embeddings [16] to replace absolute one-dimensional time differences. Since time embeddings have little impact on improving model performance in our experiment, we discuss details of this approach in **S1 Text**.

#### 4.3.1 Missing Data Handling

Our model considered 12 common laboratory tests as features, but some tests were not conducted in the same time window. The reason is that no patient has all labs drawn at the same time. We treated these unmeasured laboratory tests as missing values, which are denoted as *v*_*t*_ = 0. Previous work [17] shows that deep learning models, such as long-short term memory (LSTM) networks, can handle missing data by integrating an additional indicator *o*_*t*_ =0 for these missing values *v*_*t*_ = 0. Thus, our model used two vectors (*o*_*t*_, *v*_*t*_) instead of only one vector of observed lab test values so that the model knew which observations were missing. This so-called zero imputation strategy can handle missing data implicitly by considering feature correlations.

#### 4.3.2 Random Mask Corruption

In the training stage, the random mask corruption was designed to simulate the impact of lab test reduction on future predictions. Assuming a lab test is reduced at time *t*, and therefore, its observed value cannot be used for predicting labs at time *t* + 1. Following an earlier work[11], we randomly corrupted 10% of observed inputs (*o*_*t*_ = 1), that is, making their value *v*_*t*_ =0 at future predictions to simulate the impact of lab test reductions. We still considered the prediction errors of these corrupted Hgb values from time *t* − 1 to *t*, but did not use them to make future predictions. At the testing stage, we did not introduce any corruption and simply changed *v*_*t*_ =0 for lab tests that were recommended for reduction with respect to future predictions.

### 4.4 Development of Confidence-based Deep Learning Approach

#### 4.4.1 Feed-forward Attention LSTM

A time-aware attention mechanism [18] was used to extract essential features from the input sequence over time. First, the LSTM layer generated a sequence of hidden vectors *h*. Second, for each timestep *t*, the learnable function *a* computed the hidden vector *h*_*t*_, then produced an encoded embedding *e*_*t*_. We computed a probability weight *a*_*t*_ using a softmax function over the entire time sequence. Finally, the context vector *c* was computed as a weighted sum of the hidden vectors *h*_*t*_.

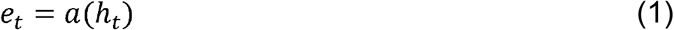

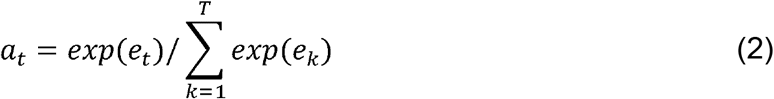

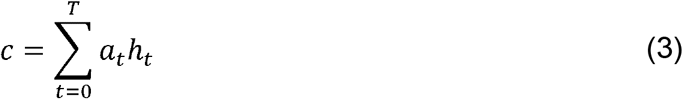

As a result, the attention mechanism enabled the model to distribute weights over the entire time period, which smoothly adjusted the impacts of long-time memories for irregular time sequences.

#### 4.4.2 Model Architecture

We presented the confidence-based deep learning work in combination with long-short term memory (LSTM) and the selective neural network under multitask learning strategies. The idea is to select a proportion of Hgb candidates with high prediction confidence so that the selected Hgb candidates are more likely to make correct classifications of normality and stability.

The architecture of our model is shown in **Fig 5**. The input features were first fed into the LSTM network module. Inspired by a multitask learning architecture[19], we used a common feed-forward LSTM layer to capture the shared information for downstream networks, and assigned two separate attention-based LSTMs. The shared LSTM layer accepted all features (i.e., laboratory values, vital signs, patient demographics, Hgb value changes, time differences, and observation indicators) as inputs, and optimized hidden states at every timestep. Demographic features were copied and placed in every timestamp. The model used two attention-based LSTM layers with shared parameters to capture task information. In the attention-based LSTM layer, input embeddings were augmented by concatenating the hidden features of the shared LSTM layer and original data *X*. One attention-based LSTM layer was followed by the stability predictor. We used Hgb values to measure the stability of future Hgb tests, and this first layer learned a subset of features, including Hgb value changes, patient demographics, time differences, and observation indicators. The other attention-based LSTM layer was followed by the normality, value, and selection predictor. These three prediction tasks require complex features and may influence each other[10]. This second layer learned entire feature vectors, including laboratory values, vital signs, patient demographics, Hgb value changes, time differences, and observation indicators.

**Fig 5.**
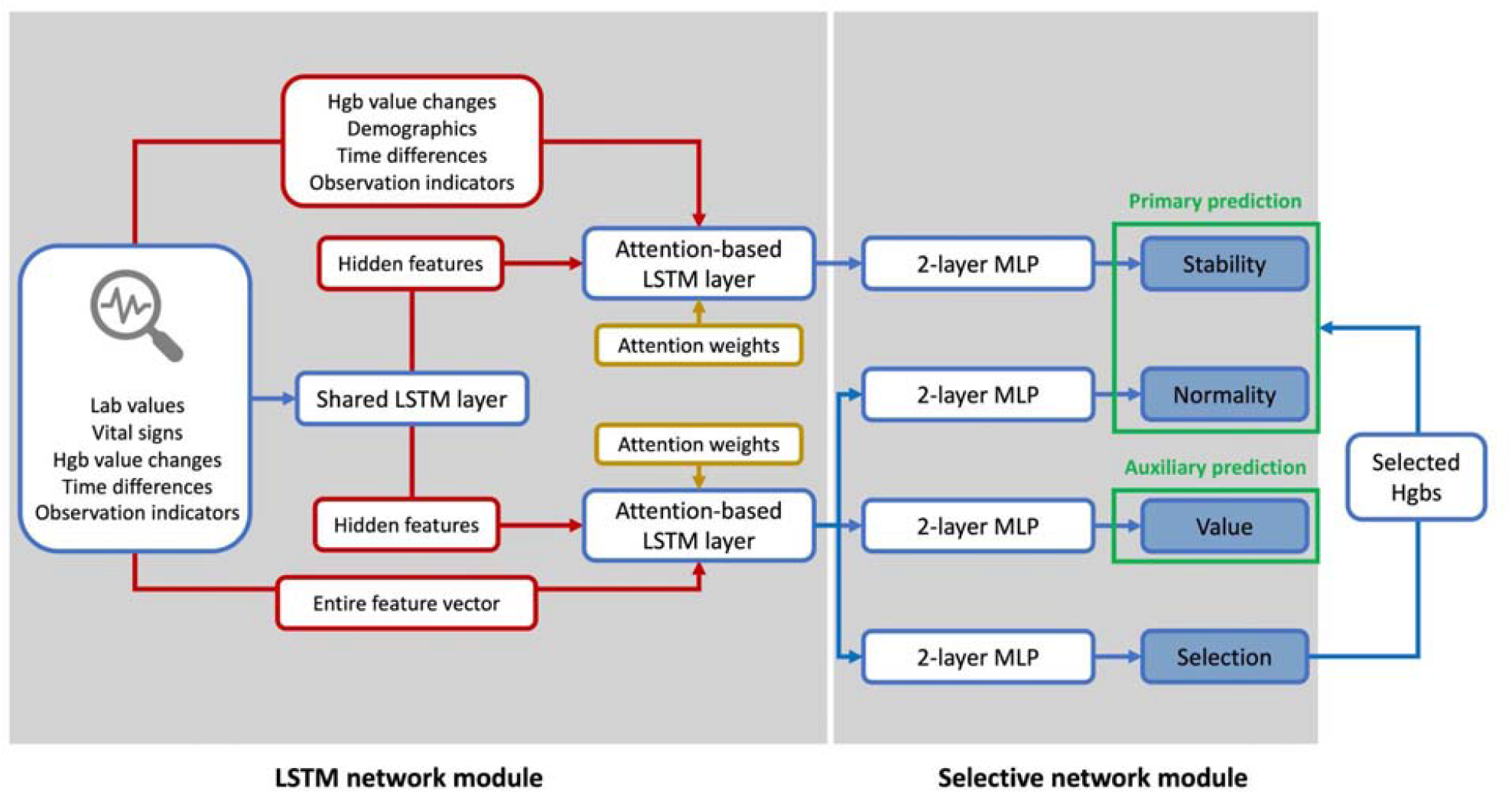
Model Architecture Framework. In the LSTM network module, the shared LSTM layer received all input features, and outputs hidden features that contained general information derived from original data. The attention-based LSTM layer augmented input embeddings by concatenating hidden features and duplicated original features. One attention-based layer learned a subset of features for the following stability predictor. The other attention-based layer learned entire feature vectors to obtain complicated information for the following normality, value, and selection predictor. In the selective network module, we had four 2-layer MLP predictors to make task-specific predictions for Hgb stability, Hgb normality, Hgb value, and selection probability in parallel. Stability and normality predictors were treated as primary predictions that focus on selected Hgb samples. The value predictor served as the auxiliary prediction that covers all Hgb samples, including the non-selected ones.

The downstream selective network module used multilayer perceptrons (MLPs) with ReLU activation, each followed by a final task-specific prediction. Each MLP predictor consisted of two fully-connected layers. The selection MLP predictor estimated the likelihood of choosing the Hgb test as a high-confidence sample[20]. More details of confidence-based selection will be introduced in the next section. The normality and stability MLP predictors were responsible for primary predictions for selected Hgb candidates. The value MLP predictor made the auxiliary prediction that covered all Hgb samples, including Hgb tests that were not selected by the model. The reason for introducing the auxiliary prediction was to avoid overfitting (i.e., the model chooses non-representative samples to only benefit normality and stability prediction). In addition, Hgb value optimization used the entire dataset to evaluate the loss in order to ensure the generalizability of normality and stability predictors.

#### 4.4.3 Loss Function with Reject Optimization

We considered the problem of selective prediction in the laboratory reduction network and leveraged the integrated reject mechanism developed in SelectiveNet[14]. It is a deep learning architecture that is optimized for selecting samples that maximally benefit model predictions. Based on our assumption that the dataset contains a proportion of Hgb outliers, such a specialized rejection model only considers a proportion of samples and filters out low-confidence ones. The approach proposes a loss function that enforces the coverage constraint using a variant of the Interior Point Method (IPM)[21]. The selection head outputs a single probabilistic value *p*_*t*_ using a sigmoid activation. At a certain timestep *t*, the selective network module achieves LSTM hidden features, then predicts Hgb normality *f*_*Y*_(*x*_*t*_) and stability *f*_*z*_(*x*_*t*_) if and only if the selection probability *p*_*t*_ exceeds a user-defined threshold *τ* (defined in **Table 4**); otherwise, the model rejects the prediction tasks of normality and stability for *x*_*t*_. Given the selection loss *L*_*f*_ in Equation (4), the performance of the selective algorithm is measured by the likelihood of normality *r*(*f*_*Y*_), the likelihood of stability *r*(*f*_*z*_), and a quadratic penalty function, *ψ*(*a*).

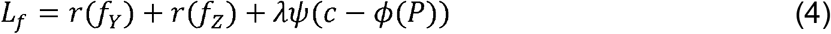

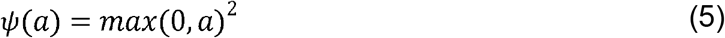

where *c* is the customized target coverage (i.e., expected proportion of samples to be considered eligible for reductions, for which the model will predict), *λ* is a penalty parameter to control the weight of the regularization. The likelihood of normality *r*(*f*_*Y*_) and the likelihood of stability *r*(*f*_*z*_) are determined by the normality loss *L*(*f*_*Y*_(*x*_*t*_), *y*_*t*_) and stability loss *L*(*f*_*z*_(*x*_*t*_), *z*_*t*_), respectively.

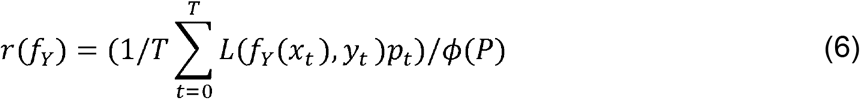

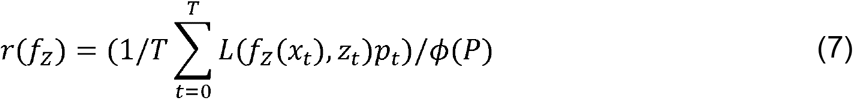

where

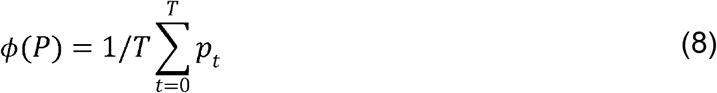

In Equation (9) and (10), the normality loss *L*(*f*_*z*_(*x*_*t*_), *z*_*t*_) and stability loss *L*(*f*_*z*_(*x*_*t*_), *z*_*t*_) are both calculated using the binary cross-entropy, where *σ* denotes the sigmoid function that converts predictions into probabilistic values.

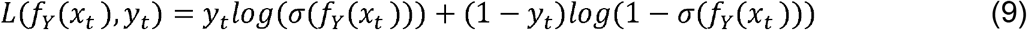

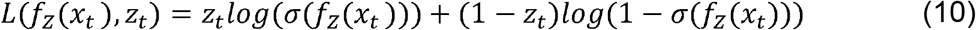

Additionally, we handled Hgb value predictions *f*_*V*_ (*x*_*t*_) as the auxiliary task using a standard mean squared error (MSE) loss function. Auxiliary predictions were used to connect the selection loss *L*_*f*_ by accounting for all samples. Otherwise, normality loss and stability loss only consider optimizing predictions of selected samples, which might cause the overfitting issue. Thus, the overall loss function *L* is a combination of the selection loss *L*_*f*_ and the auxiliary loss *L*_*aux*_ as follows:

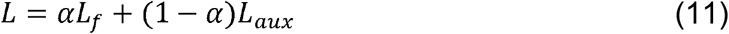

where *α* is the selection weight which lies in [0,1], and

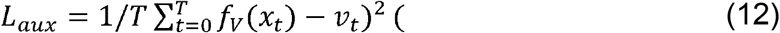

## Data Availability

These clinical data contain potentially identifying and sensitive patient information, which cannot be made available due to the privacy protection policy of the hospital.

## Ethnic Statement

Our research study was approved by the Committee for the Protection of Human Subjects (the UTHSC-H IRB) using the protocol HSC-MS-21-0452 [22]. There was a waiver of informed consent.

## Code Availability

Custom code used to analyze the model is available upon reasonable request to the corresponding author.

## Author Contributions

TH led the writing and conducted experiments. LL, EVB, XJ contributed to the generation of the idea and contributed to the writing of the paper. All authors made multiple rounds of edits and carefully reviewed the manuscript.

## Supplementary Information

### S1 Text. Supplementary Notes

#### Data analysis

Statistical analyses were conducted in Python (version 3.7). The network model is processed via PyTorch libraries using CUDA. The network was trained using the Adam optimizer, with a 64 minibatch size, an L2 regularization weight of 0.000001, and a learning rate of 0.0001 over 100 epochs. The network hyperparameters producing the lowest total loss in the test set were chosen as the final network architecture.

#### Relative Positional Time Embedding

The laboratory test data are represented as irregular time series, which have wide and heterogeneous time gaps between consecutive observations. The number of hour differences from the last laboratory draw contains positional information that indicates the relative relationships between lab test values in consequent blood draws. The time differences were concatenated with other input features (i.e., laboratory values, vital signs, patient demographics, Hgb value changes, and observation indicators) in the timestamp. Previous work [11] handled irregular time-series predictions by assigning the absolute time difference to each timestep. We assigned a unique temporal encoding to cover all possible hour gaps (relative differences by hours), which can be generalized to both short and long time gaps. Following the transformer model[16], we used sinusoidal functions to incorporate relative positions.

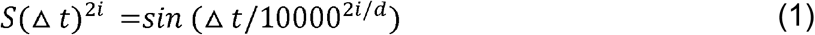

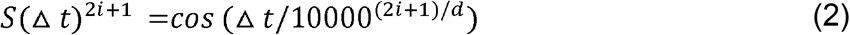

where Δ*t* is the relative time differences of laboratory tests, *d* represents the embedding dimension, and *s:N*–> *Rd* is the sinusoidal function that produces the output vector for each dimension *i*. This kind of embedding allows us to represent relative positions, in which *S*(*t* + *k*) is a linear transformation of *S*(*t*) for any fixed offset *k*.

#### Ablation Study Evaluation

To better understand the network behavior, we performed ablation experiments in which some components of the network are removed. All ablation experiments were conducted on the reduction evaluation protocol. We compared our full model with several variants with reduced structure from the full model. The model without time embeddings refers to a model considering time differences as a one-dimensional vector. The model without attention refers to a model without attention in LSTM layers. The model without confidence selection makes predictions of normality, stability, and values for all Hgb samples. The vanilla two-layer LSTM together with four two-layer MLPs whose network structure refers to Yu et al.[11]. The baseline models considering the selection mechanism (our model, without time embedding, and without attention) are set as the coverage rate at 0.85, and their corresponding model coverage rates are very close.

As shown in **S3 Table**, our full model achieves the best normality prediction performances and comparable stability prediction outcomes. Although the relative positional time embedding method works well on transformer models[16], removing time embeddings does not make a significant influence on predicting labels in our case. In our model, attention weights have improved normality AUC by 3.21% and AUPRC by 7.06%, and confidence selections have improved normality AUC by 4.98% and AUPRC by 16.13%. Compared to the vanilla two-layer LSTM network, our model includes a combination of attention weights and confidence selection, which archives performance improvements.

## Supplementary Figure

**S2 Fig.**
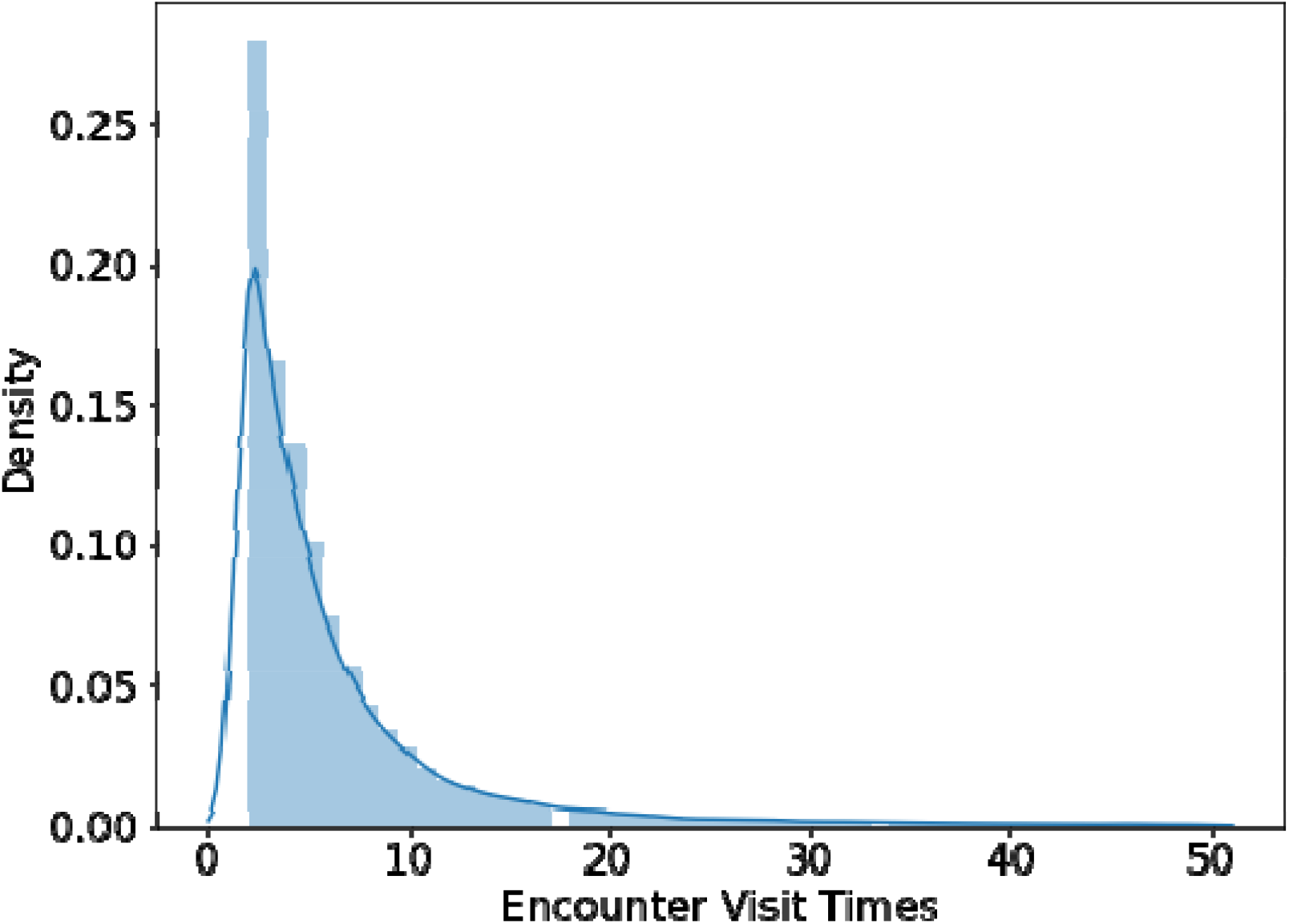
Histogram of encounter visit times. Encounter visit times refer to the number of laboratory draws for each encounter. The histogram results show that it follows a long tail distribution. Most encounters have no more than 30 laboratory test records.

## Supplementary Tables

**S3 Table.** Comparison with baseline models at an optimal coverage (*c* = o. 85). Bold numbers denote the best performances over the evaluation metric, and underlined numbers denote the second-best performance over the evaluation metric.

| Benchmark | Target Coverage | Model Coverage | Normality |  |  |  |  | Stability |  |  |  |  |
| --- | --- | --- | --- | --- | --- | --- | --- | --- | --- | --- | --- | --- |
|  |  |  | Prev | AUC | Acc | Prec | AUPRC | Prev | AUC | Acc | Prec | AUPRC |
| Our Model | 0.85 | 83.80% | 15.25% | **95.89%** | **93.17%** | 74.38% | **80.05%** | 97.40% | **95.94%** | 97.41% | 97.46% | <u>99.89%</u> |
| Without Time Embedding | 0.85 | 85.28% | 15.64% | <u>95.61%</u> | <u>92.85%</u> | **74.63%** | <u>79.53%</u> | 97.99% | 94.92% | **98.01%** | **98.01%** | <u>99.89%</u> |
| Without Attention | 0.85 | 84.75% | 15.99% | 92.68% | 90.11% | 85.53% | 72.99% | 97.95% | <u>95.44%</u> | <u>97.96%</u> | <u>97.99%</u> | **99.90%** |
| Without Confidence Selection | - | - | 20.20% | 90.91% | 85.19% | 59.28% | 63.92% | 90.90% | 94.80% | 93.48% | 95.68% | 99.46% |
| Vanilla LSTM | - | - | 20.20% | 88.40% | 82.64% | 58.10% | 61.04% | 90.90% | 94.80% | 92.91% | 94.65% | 99.47% |

**S4 Table.** Patient cohorts summarized. This table concludes input features of 3 patient demographic information (age, gender, and race), 5 vital signs, and 12 common laboratory tests.

|  | MHH Cohort |
| --- | --- |
|  | N = 62,479 |
| **Demographics** |  |
| Age, year, mean | 54.02 (0-119) |
| Male, n (%) | 25,707 (41.15%) |
| White/Caucasian | 24,032 (38.46%) |
| **Vital Signs** |  |
| Peripheral pulse rate, mean (sd) | 84.06 (18.57) |
| Respiratory rate, mean (sd) | 20.01 (5.84) |
| SpO2 percent, mean (sd) | 96.77 (3.16) |
| Diastolic blood pressure, mean (sd) | 73.47 (13.89) |
| Systolic blood pressure, mean (sd) | 130.89 (22.61) |
| **Laboratory Values** |  |
| Normal Hgb value (%) | 24.74% |
| Stable Hgb* (%) | 90.80% |
| BUN, mean (sd) | 24.42 (20.19) |
| Calcium, mean (sd) | 8.60 (0.72) |
| Chloride, mean (sd) | 105.22 (6.23) |
| Creatinine, mean (sd) | 1.56 (1.91) |
| HCO<sub>3</sub>, mean (sd) | 26.56 (7.01) |
| Hgb, mean (sd) | 11.26 (2.20) |
| Magnesium, mean (sd) | 2.15 (0.46) |
| Phosphorus, mean (sd) | 3.39 (1.25) |
| Platelet, mean (sd) | 246.71 (118.36) |
| Potassium, mean (sd) | 3.98 (0.60) |
| Sodium, mean (sd) | 138.46 (4.94) |
| WBC, mean (sd) | 9.88 (5.68) |

Sd – standard deviation. BUN – blood urea nitrogen. HCO<sub>3</sub> – sodium bicarbonate.
Hgb – hemoglobin. SpO<sub>2</sub> – percent saturation of oxygenation. WBC – white blood cell count.
\* Labs that did not change from normal to abnormal.

**S5 Table.** The number of Hgb coverages and reduction rates under reduction evaluation. This table provides numerical details of hemoglobin test reduction regarding the test dataset. The total number of encounters is 12,525. The total number of Hgb samples in the entire dataset is 57,039 starting from *t* = 0, and 44,586 starting from *t* = 1. The dominator of the coverage rate and the reduction rate is the total number of Hgbs starting from *t* = 1, which ignores initial tests at *t* = 0.

| Target Coverage | Model Coverage | Covered Hgb Counts | Reduction Rate | Reduced Hgb Counts |
| --- | --- | --- | --- | --- |
| 0.75 | 77.92% | 34,741 | 3.08% | 1,373 |
| 0.80 | 81.64% | 36,400 | 4.94% | 2,201 |
| 0.85 | 83.80% | 37,363 | 9.91% | 4,420 |
| 0.90 | 89.84% | 40,056 | 11.71% | 5,221 |
| 0.95 | 96.91% | 43,210 | 16.89% | 7,530 |
| 1.0 | 99.99% | 44,583 | 17.48% | 7,793 |

**S6 Table.** Value prediction consistency with normality under reduction evaluations. This table provides numerical details on value prediction to explain **Fig 2** in the article.

| Target Coverage | Accuracy | Accuracy within 3% | Accuracy within 4% | Accuracy within 5% | Accuracy within 10% |
| --- | --- | --- | --- | --- | --- |
|  |  | Error | Error | Error | Error |
| 0.75 | 88.32% | 95.57% | 96.78% | 97.52% | 99.63% |
| 0.80 | 89.86% | 97.97% | 99.05% | 99.47% | 99.89% |
| 0.85 | 76.15% | 93.52% | 95.91% | 97.50% | 99.86% |
| 0.90 | 65.74% | 83.24% | 87.62% | 90.94% | 99.03% |
| 0.95 | 58.49% | 79.90% | 85.30% | 89.28% | 98.40% |
| 1.0 | 50.88% | 81.09% | 87.16% | 91.22% | 99.07% |

**S7 Table.** Model performances using different selection thresholds under reduction evaluation. This evaluation table provides numerical details of selection prediction to explain **Fig 3** in the article.

| Selection Threshold | Model Coverage Rate | Normality |  |  | Stability |  |  |
| --- | --- | --- | --- | --- | --- | --- | --- |
|  |  | Prevalence | AUC | AUPRC | Prevalence | AUC | AUPRC |
| 0.05 | 85.41% | 16.64% | 95.56% | 78.36% | 96.67% | 95.31% | 99.83% |
| 0.1 | 85.04% | 16.48% | 95.67% | 78.86% | 96.84% | 95.31% | 99.84% |
| 0.15 | 84.77% | 16.38% | 95.68% | 78.85% | 96.91% | 95.44% | 99.85% |
| 0.2 | 84.55% | 16.33% | 95.80% | 79.33% | 97.05% | 95.45% | 99.86% |
| 0.25 | 84.42% | 16.23% | 95.81% | 79.32% | 97.05% | 95.51% | 99.86% |
| 0.3 | 84.26% | 16.13% | 95.82% | 79.30% | 97.07% | 95.45% | 99.86% |
| 0.35 | 84.11% | 16.09% | 95.86% | 79.49% | 97.17% | 95.52% | 99.87% |
| 0.4 | 83.99% | 15.99% | 95.88% | 79.48% | 97.19% | 95.57% | 99.87% |
| 0.45 | 83.90% | 15.96% | 95.91% | 79.61% | 97.23% | 95.62% | 99.87% |
| 0.5 | 83.81% | 15.90% | 95.94% | 79.79% | 97.30% | 95.56% | 99.87% |
| 0.55 | 83.67% | 15.82% | 95.93% | 79.78% | 97.30% | 95.66% | 99.88% |
| 0.6 | 83.53% | 15.81% | 96.00% | 80.15% | 97.37% | 95.62% | 99.88% |
| 0.65 | 83.45% | 15.73% | 96.01% | 80.11% | 97.40% | 95.71% | 99.88% |
| 0.7 | 83.28% | 15.65% | 96.05% | 80.30% | 97.47% | 95.71% | 99.89% |
| 0.75 | 83.13% | 15.59% | 96.11% | 80.60% | 97.52% | 95.70% | 99.89% |
| 0.8 | 82.97% | 15.51% | 96.12% | 80.66% | 97.60% | 95.69% | 99.89% |
| 0.85 | 82.78% | 15.40% | 96.18% | 80.97% | 97.71% | 95.75% | 99.90% |
| 0.9 | 82.52% | 15.30% | 96.24% | 81.33% | 97.82% | 95.70% | 99.90% |
| 0.95 | 82.08% | 15.06% | 96.33% | 81.75% | 97.97% | 95.87% | 99.91% |

